# The WHO Disease Outbreak News during the Covid-19 pandemic

**DOI:** 10.1101/2024.02.19.24303038

**Authors:** Ciara M. Weets, Colin J. Carlson, Hailey Robertson, Kate Toole, Lauren McGivern, Ellie Graeden, Rebecca Katz

**Author notes:** These authors share lead author status (Ciara M. Weets, Colin J. Carlson).

## Abstract

During the Covid-19 pandemic, the World Health Organization (WHO) was faced with the task of regular public updating—about both the pandemic itself, and hundreds or potentially thousands of other health emergencies. Here, we examined the 242 reports published in the WHO Disease Outbreak News (DON) during the first four years of the Covid-19 pandemic (2020 to 2023), and document the diseases and regions that were reported. We find that multinational epidemics of diseases like Ebola virus and MERS-CoV continue to dominate the DON. However, recent years have also seen more reports of climate-sensitive infectious diseases, as well as a state shift in influenza outbreak reporting in both China and the rest of the world. Surprisingly, the DON was only minimally used to document the Covid-19 pandemic and the global mpox epidemic, almost exclusively before the declaration of a public health emergency of international concern. Notably, inconsistent reporting related to Covid-19 variants of concern speaks to the ongoing evolution of the DON as a resource, and potentially, to its complicated relationship with international travel and trade restrictions. We suggest that researchers should continue to exercise caution when treating the DON as a global record of outbreak history, but that the DON is a compelling record of the WHO itself, including the process it uses to assess outbreak risk.

## Introduction

Timely notification of infectious disease outbreaks is one of the primary determinants of rapid response, and as such, is consistently one of the strongest predictors of outbreak containment and severity (Chan et al. 2010). For outbreaks of potentially high-consequence pathogens, including those of unknown origin or with known epidemic potential, the international notification process becomes particularly relevant. Under Article 6 of the International Health Regulations (IHR) (2005), when outbreaks meet certain criteria (outlined in Annex 2), Member States are required to notify the World Health Organization (WHO); under Article 9, the WHO is also empowered to collect information from other sources such as news outlets or voluntary reports from member States. The WHO then synthesizes all of this information and, in some cases, shares information directly with Member States or – in some cases – with the public.

In these cases, the most frequently used outlet (and the only official public record of outbreak history curated by the WHO) is the Disease Outbreak News (DON), an online resource that has been curated since 1996 and captures thousands of outbreak reports from around the world. Reports in the DON frequently capture the outbreak location, the causative agent (if known) or symptomology, and other critical information, such as case counts, details on response efforts, or WHO guidance. Between January 1996 and December 2019, a total of 2,789 reports were published in the DON, capturing developments in major epidemics like the 2014 West African Ebola epidemic, unusual events detected by syndromic surveillance, and everything in-between. The DON is generally understood to be an incomplete record of infectious disease outbreaks: the diseases and countries that are represented have shifted through time, reflecting not just the global history of outbreaks, but also the priorities of the WHO and the concerns of Member States. Over the last two decades, the DON has become dominated by major epidemics (such as MERS-CoV or Ebola virus outbreaks), while reports related to infectious disease of poverty (e.g. cholera) or newly emerged infections have become more sparse (Carlson et al. 2023).

Here, we revisit these trends, and explore how the WHO has used this resource in light of the unprecedented Covid-19 pandemic. We develop an incremental update to an existing dataset capturing the focus of these reports (Carlson et al. 2023), with 242 new records from between 2020 and 2023. We explore how these reports documented the Covid-19 pandemic, as well as other major epidemics and smaller outbreaks during the same time frame.

## Methods

### The Disease outbreak News

Previously, we developed a standardized dataset capturing basic information related to, and the topics covered in, the WHO Disease Outbreak News between 1996 and 2019. Here, we expand that database during the four years of the Covid-19 public health emergency of international concern (PHEIC) (2020-2023), using an updated and simplified data standard.

We reviewed all 242 reports published between January 1, 2020 and December 31, 2023 on the WHO website (www.who.int/emergencies/disease-outbreak-news). For each report, we captured the unique URL of the post (*Link*), the report title (*Headline*), and publication date (*ReportDate*). We captured outbreak geography at the country level (*Country*, as well as 3-letter ISO codes: *ISO*) or above. In reports on multi-country outbreaks within fewer than ten named countries, each report-disease-country combination was entered as a separate row. In some cases, countries were left unspecified and ISO codes were left blank if reports had an explicitly regional (e.g., “Americas”) or global focus. In keeping with the original data collection effort, we recorded a standardized set of disease names based on the language most commonly used in the reports (rather than any external taxonomy such as the International Classification of Diseases). All outbreaks are classified to the first level (*DiseaseLevel1*; e.g., “Polio”), and occasionally pathogen strains or similar granular data are also recorded (*DiseaseLevel2*; e.g., cVDPV2 or WPV:WPV1). Events of unknown etiology captured through syndromic surveillance are systematically categorized within one of seven “Syndromic” disease types (cardiovascular, diarrhoeal, gastrointestinal, haemorrhagic, hepatological, neurological, and respiratory).

### Case Data

Covid-19 case data used in Figure 2 was taken from the publicly-available COVID-19 Data Repository published by the Center for Systems Science and Engineering (CSSE) at Johns Hopkins University (Dong, Du, and Gardner 2020). Mpox case data was taken from the publicly-available Global.Health repository (Kraemer et al. 2022).

### Code and Data Availability

The updated dataset, and all code to reproduce our analyses (all conducted in R v4.3.1), are all available in a public GitHub repository (github.com/cghss/dons2).

## Results

During the four year period in which the Covid-19 PHEIC declaration was in place (2020 to 2023), the World Health Organization published a total of 242 reports in the Disease Outbreak News, describing outbreaks of 42 diseases in 89 countries (Figure 1).

**Figure 1.**
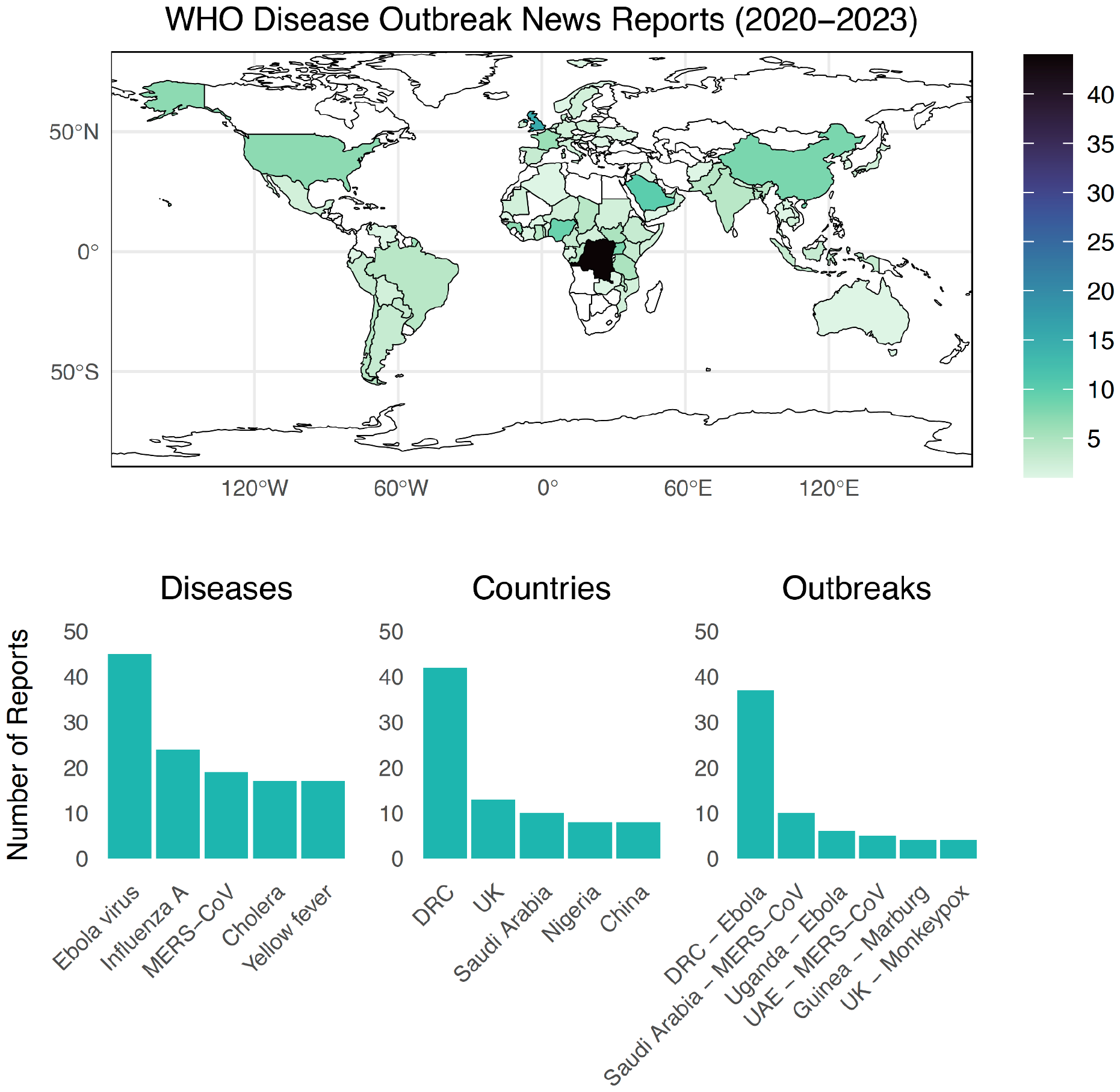
The global distribution of outbreak reporting and subject matter captured in the DON during the first four years of the Covid-19 pandemic.

### Covid-19 in the DON

Reports related to the Covid-19 pandemic accounted for a small proportion (3%) of total outbreak reporting between 2020 and 2023: only 9 reports were published related to Covid-19, all in 2020. An initial cluster of five reports were published between January 5, 2020 and January 17, 2020. The first report, on January 5 – six days after the first ProMed-mail report on December 30, 2019 – noted a pneumonia outbreak of unknown etiology in China, which the WHO China Country Office had become aware of on December 31, 2019. Notably, this report included some of the first public information related to connections between initial case clusters and Huanan Seafood Wholesale Market. Subsequent reports provided a detailed update on the situation in China, including identification of the novel coronavirus (January 12); and updates on imported cases in Thailand, Japan, and South Korea (January 14, 16, 17, and 21). On January 21, the WHO also began posting daily situation reports on COVID-19 (instead of using the DON); this continued until August 16, 2020, when reports transitioned to a weekly basis.

Covid-19 did not reappear in the Disease Outbreak News again until November 6, 2020, when a report described a novel variant that had emerged on mink farms in Denmark, which had been transmitted from mink to humans, and had acquired mutations with an unknown level of associated risk; an update on this situation was published on December 3, 2020. Another novel variant of concern that had begun to spread rapidly in the United Kingdom was noted on December 12, 2020 (“SARS-CoV-2 VUI 202012/01,” now referred to as the Alpha variant). A final report was published on December 31, 2020, providing a global update of the first year of the pandemic, including references to the variants of concern that had been recently detected in Denmark, the United Kingdom, and South Africa (“501Y.V2,” now Beta), and the ongoing challenge of genomic surveillance. At the time of writing, this is the last time that the Disease Outbreak News was used to provide updates on Covid-19. Notably, no reports were published in response to the Omicron variant, which was significantly more transmissible than all other previous variants, and drove a global spike in cases, hospitalizations, and deaths (Figure 2).

**Figure 2.**
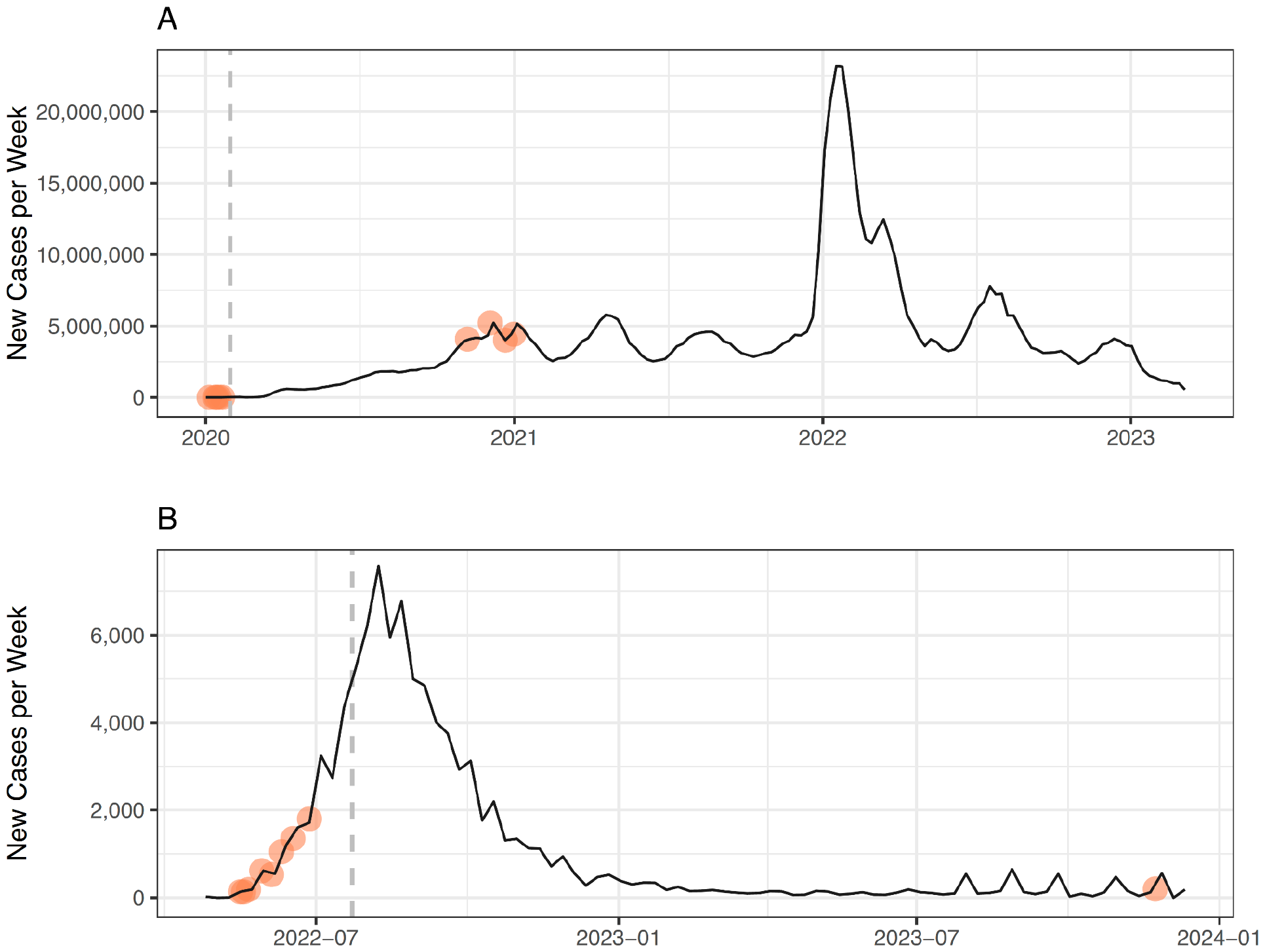
Weekly case counts and the timing of DON reports related to (A) the Covid-19 pandemic, and (B) the global mpox epidemic. Transparent orange circles indicate points where a report about the outbreak was published in the DON. Dashed lines indicate the date of PHEIC declaration by the WHO Director-General.

### Non-Pandemic Outbreaks in the DON

A total of 42 different diseases, including SARS-CoV-2, were reported in Disease Outbreak News between 2020 and 2023. During this period, the DON captured several multinational epidemics and situations, including the global mpox epidemic, multiple outbreaks of haemorrhagic fevers, and the global avian influenza (A/H5N1) panzootic.

Viral haemorrhagic fevers dominated the Disease Outbreak News between 2020 and 2023. Whereas Ebola virus was the third-most reported disease between 1996 and 2019 (Carlson et al. 2023), it was the most reported disease between 2020 and 2023, accounting for 17% of all reports (45 of 258). Most of these reports (64%) were related to the 2018 outbreak originating in the Democratic Republic of the Congo, which was declared a PHEIC on July 17, 2019 (nearly a year into the epidemic); the declaration was lifted on June 26, 2020. Even after this outbreak ended, Ebola virus remained the most reported disease in 2021 and 2022, and Marburg virus was the most reported disease in 2023. Lassa fever also appeared in 6 reports, including a February 9, 2022 report of an imported case in the United Kingdom.

The global epidemic of mpox was the first (and so far only) PHEIC declared by the World Health Organization since the start of Covid-19, and was almost entirely concurrent with the Covid-19 emergency, with the two emergency declarations being lifted a week apart. However, like the Covid-19 pandemic, this mpox epidemic only featured minorly in the DON. A total of 14 reports related to mpox were published by 2023, with all but one preceding the emergency declaration on July 23, 2022 (Figure 1B). The first report, published on October 1, 2020, described an outbreak of over 4,000 cases in the Democratic Republic of the Congo; while mpox is endemic in central and west Africa, the outbreak was characterized by an abnormally high case-fatality rate (4.2%) in children under five. In 2021, a total of four reports described imported cases of mpox that had been detected in the United States and United Kingdom, but these are believed to have been contained. Between May 16, 2022 and June 27, 2022, a series of eight reports described the emergence of an epidemic situation in high-income countries, primarily experienced by communities of men who have sex with men. On July 6, 2022, the World Health Organization transitioned to weekly situation reports instead of updates in the DON. A final report was published on November 27, 2023, drawing attention to the unprecedented and rapidly expanding outbreak of mpox in the Democratic Republic of the Congo.

Reporting patterns related to influenza shifted notably during the four year period. Historically, influenza A has been the most reported pathogen in the Disease Outbreak News, with reports from China being the most common country-disease pair. During the Covid-19 pandemic, public reporting from the World Health Organization related to health emergencies in China appears to have declined; out of 8 total reports specific to China, just 3 were related to influenza A cases. However, influenza A remained the second-most reported pathogen, with a number of reports of spillover from countries around the world—particularly related to the ongoing avian influenza (A/H5N1) panzootic. However, a handful of novel subtypes were also reported in the DONs, such as H5N8 in Russia (2020), H10N3 in China (2021), and H3N8 in China (2022), suggesting that these countries continued to comply with their obligation to report human infections with novel influenza A subtypes under the International Health Regulations (2005). One explanation for reduced reporting might be that the Covid-19 pandemic has made it much harder to distinguish causes of influenza-like illness, especially based on syndromic surveillance alone.

Finally, the last four years have witnessed a notable increase in outbreaks of climate-sensitive infectious diseases, especially vector-borne diseases such as yellow fever (25 reports) and dengue fever (14 reports), and water-borne diseases such as cholera (17 reports). These trends represent a significant state-shift in the global distribution of outbreak intensity, and the growing frequency of these reports speaks to that shift, as well as to the World Health Organization’s growing recognition of the health risks related to climate change.

## Discussion

During the Covid-19 pandemic, the international community (and the WHO) was forced to respond to an unprecedented number of concurrent health emergencies. The WHO has several means by which they share information with both member States and the public, including the Disease Outbreak News, the *Weekly Epidemiological Record*, situation reports, press releases, social media posts, and non-public communications. The question of why specific information is shared in any of these channels often speaks to the priorities of the organization as a whole, as well as the specific teams that curate each resource (Carlson et al. 2023).

During the pandemic, the WHO launched a new web platform for their health emergencies programme. The entire text of the Disease Outbreak News is now searchable, and the site includes language about the criteria used to decide which outbreaks are covered, which closely mirrors the decision framework used to identify possible PHEICs under the IHR (2005) (“Disease Outbreak News (DONs)”, 2024):

*Disease Outbreak News (DONs) are published relating to confirmed or potential public health events, of:*

- *Unknown cause with a significant or potential international health concern that may affect international travel or trade;*
- *A known cause which has demonstrated the ability to cause serious public health impact and spread internationally;*
- *High public concern which may lead to disruption of required public health interventions, or could disrupt international travel or trade*.

These changes represent a significant advance in the level of transparency and care with which the Disease Outbreak News is handled: historically, the criteria for reporting have been opaque and inconsistent, with limited searchability and even outright loss of some reports during system transitions (Carlson et al. 2023). These changes also help clarify the unique value proposition of the DONs compared to, say, the all-purpose outbreak reporting in sources like ProMed-mail and HealthMap: they represent a first-order, qualitative risk assessment by the WHO related to which outbreaks have become, or are likely to become, emergencies.

As the WHO becomes clearer about the purpose of the DON, the DON itself becomes a more revealing record of the WHO’s internal process. Some “usual suspects” – like novel subtypes of influenza, or viral haemorrhagic fever outbreaks – will always meet the criteria described above, and are likely to continue to be publicized through routine channels. The growing representation of climate-sensitive diseases similarly speaks to WHO’s recognition of their growing potential to cause disruptions to travel and trade in Africa and Latin America especially. The most interesting cases may be where WHO breaks from this established process: for example, the Alpha and Beta variants of Covid-19 were recorded in the DON, but the Omicron variant, which not only had a higher impact on human health, but also led to more significant international disruptions of travel and trade, was not. Decisions like these may simply be stochastic and may be reflective of high-consequence, limited-time frame decision-making, but they may also sometimes reflect the challenges faced by the WHO as global governance changes. For example, the Omicron variant led to an unprecedented breakdown of international norms around travel restrictions, which now poses an enduring problem in future outbreaks (Schermerhorn et al. 2022). It remains to be seen how these new challenges will change the baseline process of information sharing during health emergencies—making the DON a more important resource than ever.

